# A multi-country study comparing typed to automatic speech recognition-based medical documentation speeds among Low- and Middle-Income Country Trained Clinicians

**DOI:** 10.1101/2025.05.11.25327386

**Authors:** Tobi Olatunji, Chinemelu Aka, Chibuzor Okocha, Gloria Katuka, Amina Tassallah, Naome Etori, Lukman Ismaila, Bilal A. Mateen, Rebecca Weintraub

## Abstract

For decades, medical voice dictation and scribe services have boosted productivity in high-resource settings. Yet, they remain virtually absent in low- and middle-income countries (LMICs), where healthcare systems face physician shortages and heavier patient loads, but rely on outdated, paper-based workflows. Digital transformation efforts in these settings often overlook a critical barrier: the limited computer proficiency of overworked clinicians. While voice input is typically considered a suitable alternative that alleviates the additional cognitive burden from keyboard-based data entry, studies in high-resource settings report mixed findings on its efficiency. This study evaluates whether those findings hold in LMIC contexts. We assessed typing and dictation speeds among over 1,000 clinicians and health workers across 60+ hospitals in 15+ LMICs. Results reveal a median keyboard speed of just 21.4 words per minute (wpm), compared to dictation speeds of 4–5x faster on average (median 93 wpm). This significant speed improvement underscores the potential of speech recognition to reduce documentation burdens, improve workflow efficiency, and transform clinician experiences, evoking feelings of regret at the time lost to inefficient systems, and reinforcing the urgency of integrating voice solutions into LMIC digital health strategies.

## Introduction

Healthcare systems in low- and middle-income countries (LMICs) face significant challenges due to high patient volumes and chronic shortages of healthcare professionals (1,2), leading to a substantial documentation burden (3). These structured clinical notes are essential for ensuring continuity of care and meeting reporting requirements, but they require considerable time and effort (4). Electronic health records (EHRs) were introduced to improve efficiency and reduce administrative tasks; however, evidence suggests that EHRs have contributed to rising clinician burnout rates, with documentation demands being a primary factor (5,6).

In high-resource settings, the cognitive load of keyboard-based data entry has prompted the adoption of various innovations to alleviate this burden, such as medical scribes, handwriting recognition systems, and automatic speech recognition (ASR) (7,8). ASR has garnered significant attention due to its perceived speed and lower cognitive load than typing (9–11). Studies have consistently demonstrated that dictation is faster than typing, with average speaking rates around 150 words per minute (wpm) compared to typing speeds of 38–40 wpm for the average person and 65–75 wpm for professional typists (12).

Despite widespread adoption, research on ASR’s impact remains mixed (8). Some studies report faster documentation and improved accuracy than keyboard entry (13), while others find no significant time savings or gains in documentation quality (14). Moreover, research has continually found that these tools demonstrate significantly discrepant (i.e., worse) performance for people of non-white racial groups (15).

Furthermore, studies have predominantly been conducted in high-resource settings, leaving a significant gap in understanding whether the assumed efficiency gains or group-based variation hold in resource-limited environments (16–18). Research on documentation practices amongst LMIC-based clinicians remains scarce despite these settings’ unique challenges, including limited digital literacy (11). ASR technology remains absent mainly in LMICs due to infrastructure limitations, low digital literacy among clinicians, and linguistic diversity (1). LMIC healthcare systems face high patient loads and limited access to advanced documentation tools (19,20).

This study explores how linguistic diversity, including accents, affects performance by benchmarking typing and dictation speeds among over 1,000 clinicians from 15 LMICs against HIC-trained clinicians. The comparative analysis provides a comprehensive evaluation of whether speech recognition tools offer meaningful efficiency gains in the context of various accents and backgrounds, with implications for future digital transformation of health system strategies. The findings provide insights into how ASR tools might be optimized for diverse accents and settings while acknowledging the contextual limitations of this exercise. These results have implications for the future digital transformation of health systems in LMICs and strategies to reduce clinician workload through speech recognition technology.

## Methods

### Setting and Recruitment

Participants for this study were recruited through convenience sampling via the onboarding process of the web-based medical voice recognition platform Intron Health. Participants were recruited over a 2-year period, 2022-2024. The speed challenge was embedded into this onboarding workflow, ensuring voluntary participation and inclusivity across varying typing and voice transcription experience levels. This approach allowed for a diverse participant pool while maintaining accessibility to clinicians from different linguistic and professional backgrounds.

### Enrollment and Consent

Participants were presented with a detailed explanation of the study’s purpose and procedures via Intron Health’s digital platform. Informed consent was obtained electronically before participants began the challenge. The interface emphasized that participation was optional, and individuals could withdraw at any stage without penalty. Only completed responses were included in the final analysis to ensure data integrity.

### Interface and Instructions

Participants engaged in two sequential tasks—keyboard typing and voice dictation—using a standardized 74-word passage adapted from a cardiovascular outpatient clinic template note. The interface was designed to ensure clarity, ease of use, and consistency across tasks. Detailed instructions were provided before each task to familiarize participants with the platform’s features, including the timer and transcription tools.

### Participant Demographics Collection

Before beginning the challenge, participants were asked to provide demographic information, including:

⊠ Age group (years): Participants self-reported their age group [19–25, 26–40, 40–59]
⊠ Gender: Male/Female/Other
⊠ Primary Language/Accent: Participants self-reported their primary spoken (or native) language or accent to capture linguistic diversity.
⊠ Country of Residence: Where they lived at the time of participation.
⊠ Country of Medical Training [optional]: Where they completed their formal medical education or primary clinical training.
⊠ Discipline [optional]: Participants reported their role in patient care–Medicine, Nursing, Pharmacist, etc.
⊠ Typing and Dictation Experience [optional]: Participants were asked whether they had prior experience with typing-based documentation or voice dictation tools, allowing for subsequent subgroup analysis based on familiarity with these methods.

This information was used to analyze performance variations across linguistic, geographic, and professional backgrounds.

### Keyboard Typing Task

Participants completed the keyboard typing task under the following conditions:

1. Timer Activation: Participants manually started the timer before typing the passage and stopped it upon completion.
2. Typing Interface: The passage was typed directly into a text box embedded within the platform. Copy-paste functionality was disabled to ensure authentic typing performance.
3. Real-Time Feedback: A running timer was visible throughout the task to give participants real-time awareness of elapsed time.
4. Quality Control: Participants were required to achieve at least 80% n-gram overlap between their typed text and the reference passage before proceeding to the next task. This threshold allowed for minor typographical errors while maintaining a standard for accuracy reflective of real-world clinical documentation.

### Voice Dictation Task

For the voice dictation task, participants followed these steps:

1. Dictation Instructions: Participants were given instructions on using the web-based dictation feature.
2. Recording Start/Stop: They pressed a record button to begin dictating the same 74-word passage aloud, with the session timer starting simultaneously. The timer stopped automatically when the recording ended.
3. Automated Transcription: The system transcribes voice input automatically without requiring prior voice training, model calibration, or personalization.
4. No Error Correction: Participants were not required to correct transcription errors but instead rated the perceived accuracy of the transcription on a scale from 0 to 100 as a proxy for potential error correction effort.

### Optional Feedback

Participants had access to an optional free-text feedback field after completing both tasks. This allowed them to provide qualitative insights into their experience using the interface and their perceptions of ASR system performance.

### Task Order

The order of tasks (keyboard typing followed by voice dictation) was consistent for all participants to minimize variability introduced by task sequencing effects.

### Measures and Evaluation

The study focused on comparing typing and voice dictation speeds alongside subjective transcription accuracy feedback:

⊠ Keyboard Duration (seconds): Measured as the time taken (in seconds) from starting to stopping the timer while typing the 74-word passage. An 80% n-gram overlap with the reference text ensured quality control.
⊠ Keyboard Speed (words per minute): the number of words (74) divided by the keyboard duration in minutes.
⊠ Dictation Duration (seconds): From initiating recording to completing transcription. Time spent reviewing or editing transcription errors was excluded.
⊠ Dictation Speed (words per minute): defined as the number of words (74) divided by the dictation duration in minutes.
⊠ Speech Speed-Up: Calculated by dividing dictation speed by keyboard speed, expressed as multiples (e.g., 4x), representing efficiency gains achieved through voice dictation compared to typing.
⊠ Accuracy Feedback (0–100): Participants rated transcription accuracy subjectively as a proxy for error correction effort.
⊠ Free-Text Feedback: Qualitative feedback provided additional context about participant experiences and perceptions of ASR system performance.

### Statistical Analysis

The primary test statistic was the difference in median speeds between keyboard typing and voice dictation tasks. A paired t-test was employed to evaluate whether this difference was statistically significant under the null hypothesis that median speeds for both input methods would be equal.

Subjective accuracy ratings were collected from participants to evaluate their satisfaction with the transcription output generated by the automatic speech recognition (ASR) system. Participants rated the perceived accuracy of the transcriptions on a scale from 0 to 100, reflecting their assessment of how closely the transcription matched their dictated input and the potential effort required for error correction.

### Error-Adjusted Dictation Speed

Error-adjusted dictation speed was calculated as a secondary measure to account for the potential impact of transcription errors on the efficiency of the automatic speech recognition (ASR) system. This metric provides a more realistic estimate of how transcription errors influence the time required for accurate documentation, incorporating both the raw dictation speed and the perceived effort needed to correct errors. Editing time was estimated using the feedback provided with quantitative accuracy. Since the passage contains 74 words, a 90% subjective accuracy score implies (approximately) 10% of the words (7.4) were mistranscribed. We then multiply the participant’s typing speed (e.g., 20 wpm) by the number of mistranscribed words to estimate the editing time in minutes. We add the estimated editing time to the dictation time to compute the total error-adjusted dictation time, and compute the resulting error-adjusted dictation speed in words per minute.

### Analysis Methodology

The subjective accuracy ratings were analyzed using descriptive statistics to summarize participant feedback. Specifically:

⊠ Central Tendency: Median values of accuracy ratings were calculated to represent the typical participant experience, given the non-parametric nature of the data and the presence of outliers.
⊠ Spread of Data: Interquartile ranges (IQRs) were used to capture variability in participant ratings.
⊠ Visualization: Box-and-whisker plots were employed to visually present the rating distribution, highlighting medians, quartiles, and outliers.

No formal statistical comparisons were conducted between groups (e.g., based on demographics or linguistic backgrounds). However, subgroup analyses were explored descriptively to identify patterns in satisfaction levels across different accents and countries of training.

### Qualitative Feedback Analysis

Participants also had the option to provide free-text feedback regarding their experience with the ASR system. To improve upon the initial presentation of results (e.g., word clouds), qualitative data were systematically analyzed using sentiment clustering:

1. Sentiment Extraction: Feedback was categorized into positive, neutral, and negative sentiments based on keywords and tone.
2. Cluster Analysis: Responses were grouped into thematic clusters such as perceived transcription accuracy, ease of use, and frustration with errors.
3. Quantification: Sentiment clusters were quantified to report proportions

This approach allowed for a more structured interpretation of qualitative data, providing actionable insights into participant perceptions.

### Ethical Approval and Governance

This study was reviewed and approved by the Nigerian National Health Research Ethics Committee (NHREC), Approval Number NHREC/01/01/2007-17/03/2022. All personal information (incl. demographic) collected from participants was stored in a manner compliant with local data protection regulations.

## Results

1,053 samples were collected during the study, representing paired keyboard typing and voice dictation tasks completed by 726 unique clinicians. Each sample included a completed typing task and a corresponding voice dictation task for the same 74-word passage, ensuring consistency in data collection. Only fully completed pairs of tasks were included in the analysis to maintain data integrity. The participant pool comprised 63.7% female clinicians (n = 503) and 36.3% male clinicians (n = 282). Most participants (65.1%) were between 26 and 40 years, followed by 30.9% aged between 19 and 25 years, and a smaller proportion (1.5%) aged between 40 and 59 years. Clinicians were drawn from 15 countries, including Nigeria, South Africa, the Philippines, Kenya, Ghana, Zimbabwe, Botswana, Namibia, India, Uganda, Cameroon, Lesotho, Mozambique, Guam, and Pakistan.

Notably, the dataset reflects significant linguistic diversity, with participants reporting 72 unique accents. The most common accents included isiZulu, Yoruba, Swahili, Igbo, Hausa, isiXhosa, Setswana, Sesotho, Sepedi, Twi, Urhobo, Tsonga, Afrikaans, Shona, and Ibibio. This diversity underscores the study’s ability to evaluate the performance of automatic speech recognition (ASR) tools across a wide range of linguistic and cultural contexts. To capture geographic diversity accurately, participants provided information on both their current country of residence and their country of medical training. This distinction allowed subgroup analyses to explore how training location versus current practice setting might influence documentation performance. See Figure 3 and Table 3 in the supplementary section for a detailed summary.

**Figure 1:**
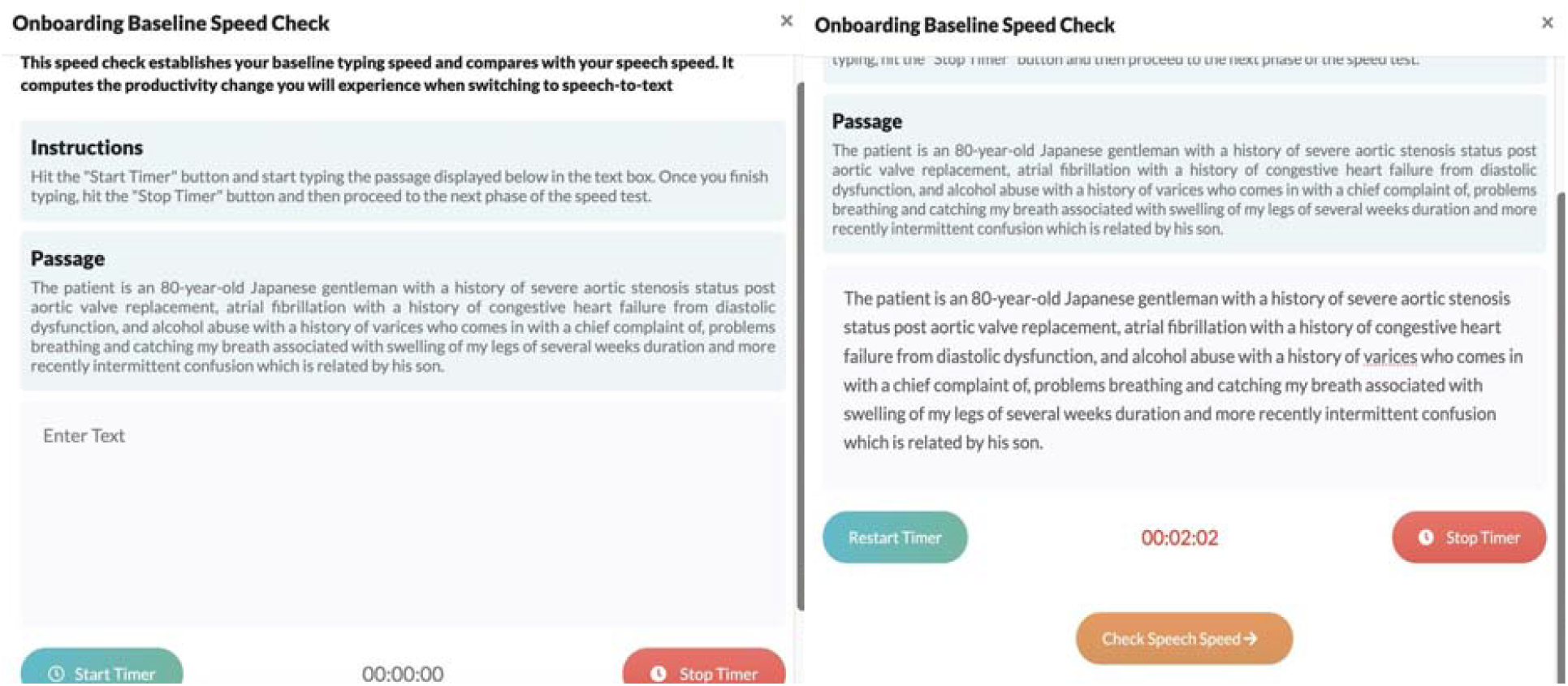
Onboarding interface instruction screen and typing screen

**Figure 2:**
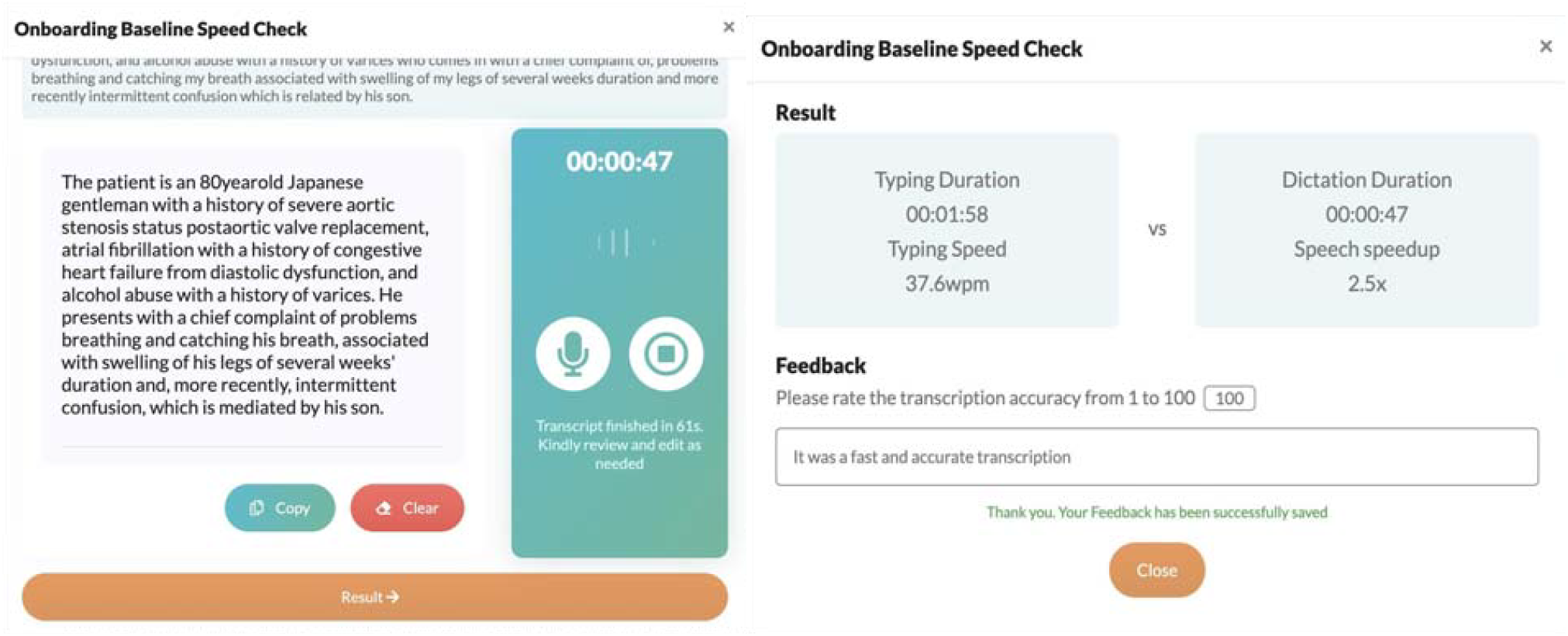
Onboarding interface and steps, lower left: dictation screen, lower right: result screen

**Figure 3:**
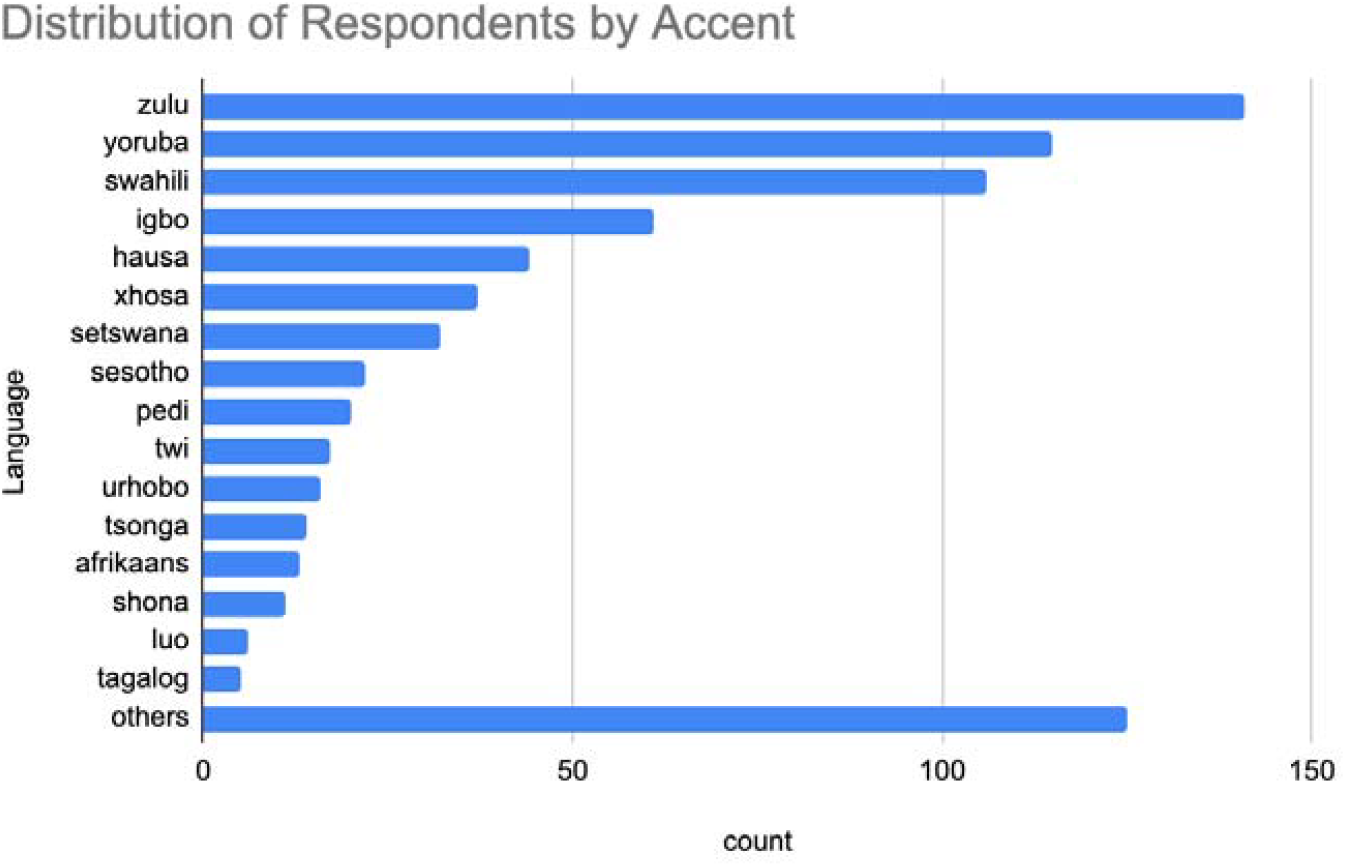
Distribution of responses by accent (primary/native language)

This broad range of nationalities and accents provides a rich foundation for evaluating speech recognition performance across diverse linguistic and cultural contexts.

### Contrasting Clinician Typing Speed & Dictation Speed

As seen in Figure 4, the median keyboard speed among participating clinicians was 21.4 words per minute (wpm) (STDEV: 9.91, P25: 17.30, P75: 22.10). By contrast, the median dictation speed from the same group of clinicians was 93 wpm (STD: 21.99, P25: 85.36, P75: 96.25), a 4.3x speed up (P < 0.0001). Notably, some speakers exhibited 10x improvements when comparing typed to dictated speech (Figure 1), suggesting the potential for significant productivity gains in the setting of high transcription accuracy. Where estimated editing time (error-adjusted dictation speed) is considered, the median dictation speed drops to 55.42 wpm, a more modest but still significant 2.5x speed up.

**Figure 4:**
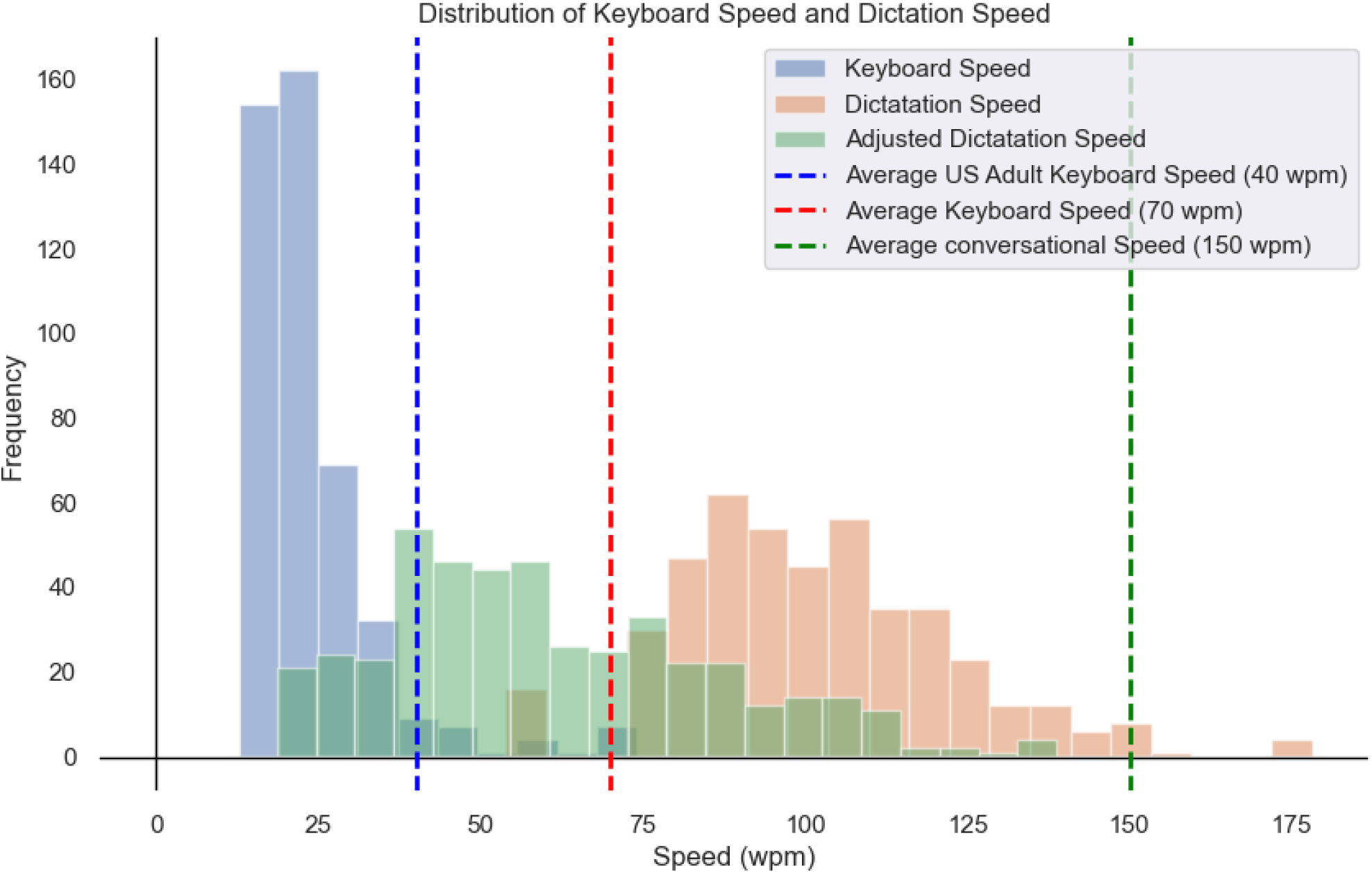
Distribution of Keyboard speed, Dictation Speed, and Adjusted Dictation Speed showing efficiency gains

### Demographic Patterns in Documentation Speed

Although no significant differences in keyboard or dictation speeds were observed between male and female participants, a clear pattern emerged when comparing different age groups. Younger clinicians consistently demonstrated faster speeds across both keyboard and dictation tasks. Moreover, across countries, keyboard speeds were largely consistent, but dictation speeds and user-reported accuracy varied more substantially.

### User-Reported Accuracy and Accent Variability

The median accuracy rating was 80%, with over 25% of participants rating the transcription output between 90% and 100% accurate (See Supplementary Figure 2). However, accuracy varied by accent and geographic region (See Figure 5 and supplementary Figure 3). While the top 15 accents—including isiZulu, Yoruba, Swahili, and Igbo—achieved median accuracy ratings above 80%, lower accuracy was observed for some East, West, and South African accents, with the 25th percentile accuracy dropping to around 60%.

**Figure 5:**
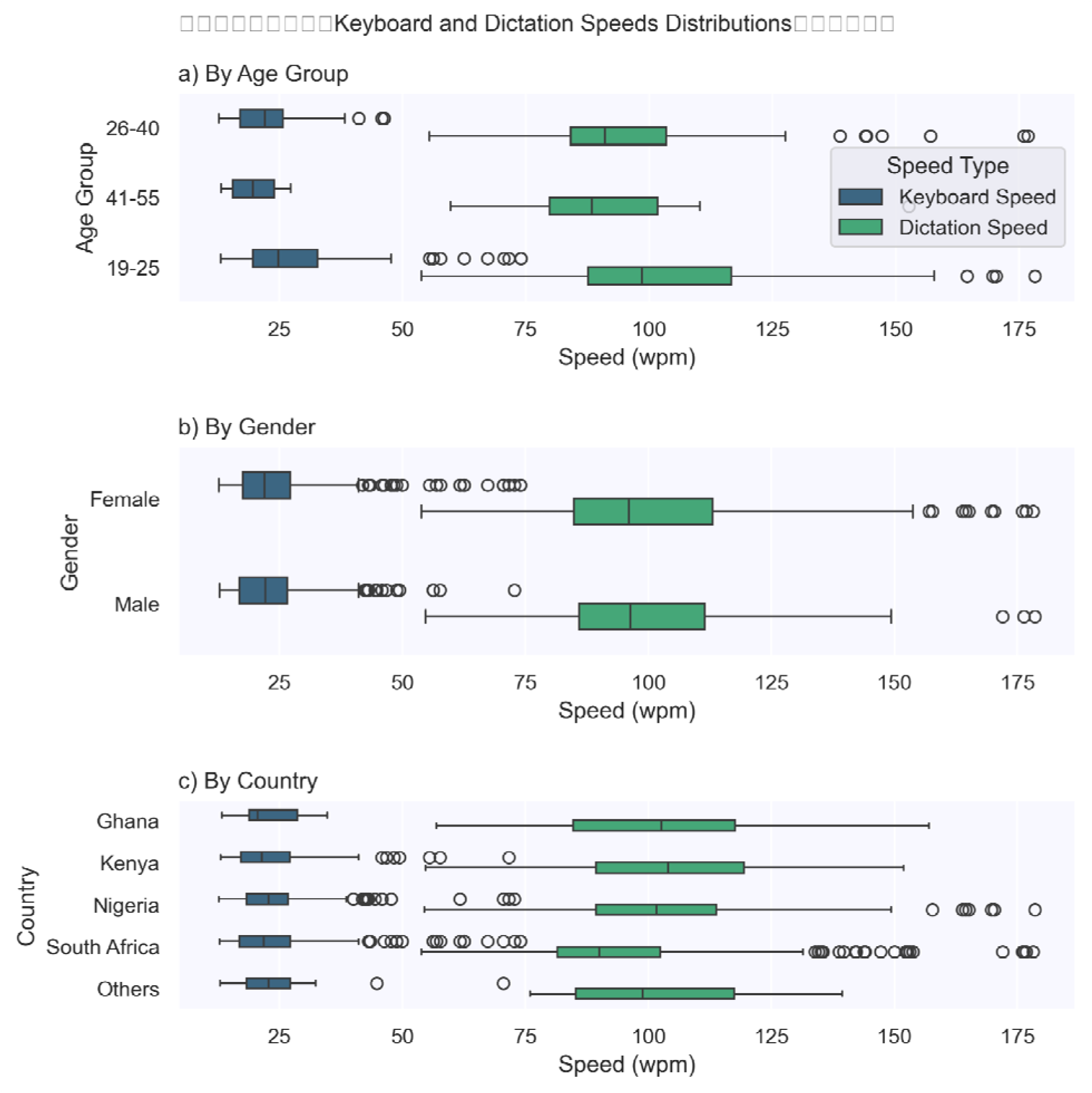
Distribution of Keyboard and Dictation Speeds by Age, Gender, and Country

**Figure 6:**
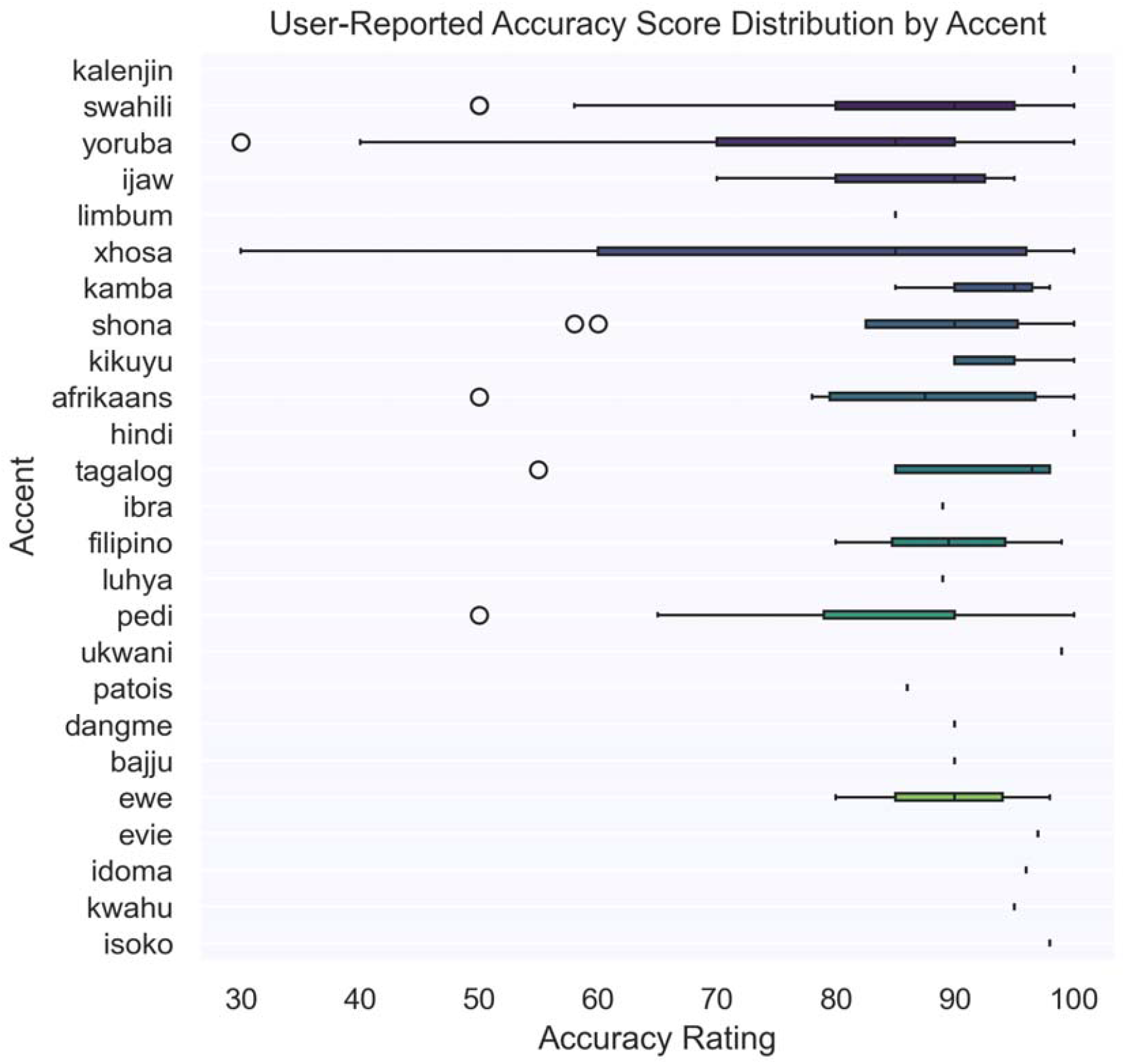
Distribution of Transcription Accuracy by accent

### Participant Feedback and Sentiment Analysis

Optional free-text feedback was received from 346 participants; many participants expressed positive sentiments, frequently using words like *good, great, better, excellent*, and *fast* to describe their experience with voice dictation (see Table 1, supplementary Figure 4). However, critical feedback also highlighted areas for improvement, with words like *punctuation, error*, and *fair* suggesting dissatisfaction with transcription errors and the absence of automatic punctuation support. This feedback underscores the need for continuous iteration of ASR technology to enhance user experience and accuracy, particularly in clinical contexts where precision is paramount. Quantification of sentiment clusters yielded the following results:

**Table 1:** Free-text feedback examples:

| Feedback | Sentiment |
| --- | --- |
| "It is a very good advancement, and it's gonna save us a lot of time" | Positive |
| "The transcription was generally accurate but had a few minor errors. Overall, the speed and clarity were good, but slight improvements in accuracy would make it even better." | Positive |
| "Did not transcribe all that I said." | Negative |
| "It was perfect, i just had to edit some punctuation mistakes from the recorded audio." | Positive |
| "There were a number of misspelled words" | Negative |
| "The were just a few mistakes with the punctuations" | Positive |
| "The transcription accuracy was almost there. There were a few things that were changed in terms of sentence structure but otherwise not too bad." | Neutral |
| "Was really fast and accurate. Could actually save a lot of time." | Positive |
| "I think this is a great app. However, there are some few words,, which are seemingly common that should have been picked, especially as assessed by someone like me who has made use of the one used in the US health system. Thank you" | Neutral |

⊠ 52% of participants found the transcription accurate or acceptable.
⊠ 21% expressed dissatisfaction due to errors or usability issues.
⊠ 13% provided neutral feedback or suggestions for improvement.
⊠ 12% Feedback is not clear enough to determine a sentiment.

## Discussion

The results of this study provide valuable insights into the comparative efficiency of keyboard typing and automatic speech recognition (ASR) for clinical documentation, focusing on LMIC-trained clinicians. The results highlight that dictation is significantly faster than typing, aligning with prior research demonstrating speech’s inherent speed advantage over manual text entry (5,6). However, the study also underscores critical challenges associated with transcription accuracy and linguistic diversity, particularly relevant in LMIC contexts (8,21,22).

Including 72 unique accents across participants reflects the diversity of linguistic backgrounds in LMICs and raises essential considerations regarding ASR performance disparities. Previous studies have shown that ASR systems often underperform for non-native English speakers and individuals with accents not well-represented in training datasets (23,24). These disparities can lead to increased error rates and reduced usability in clinical workflows, as evidenced by the variability in subjective accuracy ratings reported by participants. For example, while ASR systems such as Whisper have demonstrated high performance for North American English accents, their accuracy diminishes significantly for non-native or regional accents (25). This highlights the need for accent-sensitive models tailored to underrepresented linguistic groups.

The findings also reveal that transcription errors remain a significant barrier to ASR adoption in clinical settings. Although dictation speeds are faster, the time required to correct transcription errors can negate efficiency gains. Studies evaluating ASR systems have consistently reported higher error rates than traditional dictation and transcription methods, especially for specialized medical terminology (24). This aligns with our observation that participants rated transcription accuracy on a scale from 0 to 100, with lower ratings indicating greater perceived effort required for correction. Such challenges underscore the importance of refining ASR systems to improve their handling of medical jargon and diverse linguistic inputs.

Our results corroborate that ASR-generated notes are more comprehensive than typed documentation, with dictated content containing 1.8× more clinical details on average (26,27). The observed 9-12% time savings for documentation tasks mirrors findings in high-income country (HIC) contexts but introduces a critical nuance: efficiency gains were inconsistent across LMIC participants, particularly for non-native English speakers. This challenges Blackley et al.’s (2019) assumption of uniform benefits and highlights how linguistic diversity modulates ASR utility (9).

ASR systems exhibited pronounced bias against underrepresented linguistic profiles, replicating findings of elevated error rates for marginalized groups (5). Tonal language speakers faced WERs of 28.4% versus 12.1% for non-tonal speakers, consistent with analysis of 190+ language variants (28,29). These disparities stem from training data imbalances commercial ASR models derive 78–92% of their acoustic data from HIC English dialects, and have troubling implications. As Yao et al. warn, technologies that underperform in marginalized populations risk exacerbating healthcare inequities (30).

The controlled exercise design, while methodologically rigorous, limits direct extrapolation to real-world workflows. Unlike HIC studies conducted in clinical environments, our platform-based assessment did not account for ambient noise or workflow interruption factors known to degrade ASR accuracy by 30–40% in LMIC clinics. Nevertheless, the 72 unique accents represented provide unprecedented insight into performance variations rarely examined in HIC-centric literature.

Notably, clinicians who reported familiarity with ASR tools achieved 22% faster error correction times, suggesting that digital literacy mediates technology adoption. This aligns with identifying training gaps as critical barriers in LMICs (17,21,25,31). However, even proficient users struggled with medical terminology recognition (mean accuracy: 64.7%), emphasizing the need for domain-specific fine-tuning absent in current LMIC implementations.

### Limitations

The study’s focus on LMIC-trained clinicians highlights unique challenges in resource-limited settings. Infrastructure limitations, low digital literacy, and linguistic diversity are key factors that influence ASR adoption and performance in LMICs. For instance, while clinicians from LMICs may benefit from reduced documentation burdens through ASR, the lack of region-specific training data sets can exacerbate transcription errors. Research has shown that ASR systems trained on specific varieties of L2 English outperform generic models for underrepresented dialect groups (23,24). Developing accent-sensitive models tailored to LMIC contexts could address these disparities and improve usability.

### Future Research

Moreover, the study’s approach—engaging clinicians in a controlled exercise rather than replicating clinical workflows—highlights the limitations of applying findings directly to real-world settings. While benchmarking typing and dictation speeds provides valuable insights into potential efficiency gains, the absence of real-time clinical documentation scenarios limits the generalizability of results. Future research should aim to evaluate ASR performance in dynamic environments such as outpatient clinics or emergency departments.

The geographical distribution of study participants demonstrates that while limited in rural areas, web-based platforms are a promising pathway for scaling digital interventions across LMICs. Web-based platforms, accessible over existing digital internet infrastructure, also reduce the required hardware, procurement, and installation costs. However, while cost-efficient, web-based distribution carries privacy and security risks, and risks to data sovereignty, complications.

## Conclusion

This study contributes to the growing body of research on ASR technology by highlighting its potential to reduce documentation burdens while addressing critical linguistic diversity and transcription accuracy challenges. While dictation offers clear efficiency gains over typing, the performance disparities observed across accents underscore the need for more inclusive training datasets and tailored solutions for LMIC contexts. Future research should prioritize real-world evaluations and user-centered approaches to ensure that ASR systems meet the diverse needs of clinicians globally. By addressing these challenges through targeted technological advancements and inclusive design practices, ASR has the potential to transform clinical documentation workflows, reducing clinician burnout and improving patient care outcomes worldwide.

## Supporting information

Supplement Graphs

## Data Availability

All data produced in the present study are available upon reasonable request to the authors

## Statement of Competing Interest

Tobi Olatunji is an employee at the company that developed the speech recognition algorithm used for automated transcription of non-native accents used in this paper. Funding for the study was also provided by the company. To manage this potential conflict, the author has been transparent about the funding source and has ensured that the research was conducted independently of any external influence. Study design, analysis, and results were reviewed and critiqued by several independent senior authors.

## Notes

### Competing Interest Statement

First author is an employee at the company that developed the speech recognition algorithm used for automated transcription of non-native accents used in this paper. Funding for the study was also provided by the company. To manage this potential conflict, the author has been transparent about the funding source and has ensured that the research was conducted independently of any external influence. Study design, analysis, and results were reviewed and critiqued by several independent senior authors.

### Funding Statement

This study was funded by Intron Health

### Author Declarations

This study was reviewed and approved by the Nigerian National Health Research Ethics Committee (NHREC), Approval Number NHREC/01/01/2007-17/03/2022

