## Supplement Graphs for "A multi-country study comparing typed to automatic speech recognition-based medical documentation speeds among Low- and Middle-Income Country Trained Clinicians"

^1^ Intron Health

^2^ BioRAMP Labs

^3^ Georgia Institute of Technology

^4^ Brown University

^6^ Microsoft

^7^ University of Minnesota-Twin Cities

^8^ Johns Hopkins University

^9^ University of Florida

^10^ PATH

^11^ University of Birmingham

^12^ Harvard Medical School

**Corresponding Author**

Tobi Olatunji

Key Words: dictation, keyboard speed, speech recognition, LMIC

Word Count: 3658

Tables: 1

Figures: 6

Supplementary Material

Supplementary Fig 1: Distribution of Dictation Speed Up

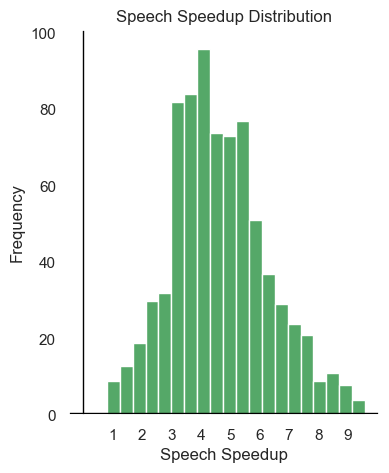

Supplementary Fig 2: Distribution of Accuracy Ratings

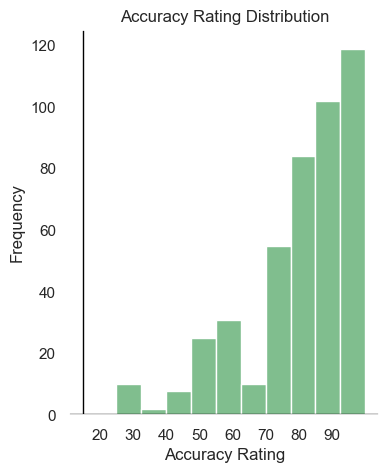

Supplementary Fig 3: Distribution of Accuracy rating by Country

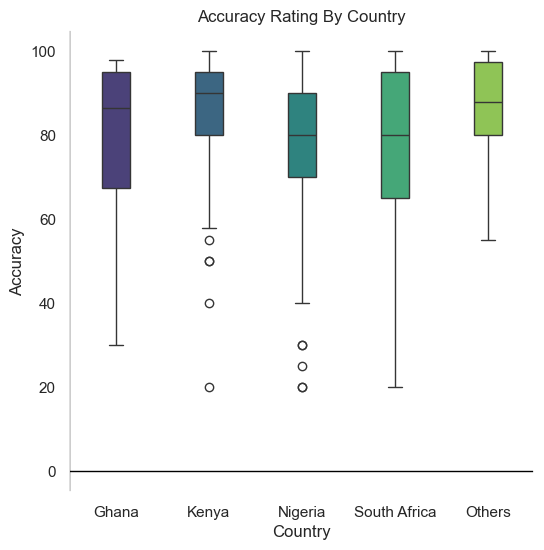

Supplementary Fig 4: Word Cloud of free text feedback

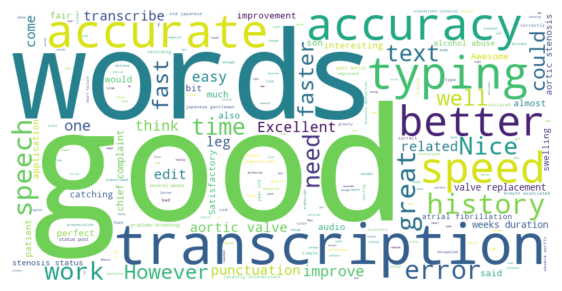

Supplementary Table 1: Country Counts

| **Country** | **count** |
| --- | --- |
| Nigeria | 313 |
| South Africa | 294 |
| Kenya | 119 |
| Ghana | 30 |
| Philippines | 8 |
| Zimbabwe | 7 |
| Botswana | 3 |
| India | 2 |
| United States | 2 |
| Cameroon | 1 |
| Uganda | 1 |
| Jamaica | 1 |
| Mozambique | 1 |
| Pakistan | 1 |
| Lesotho | 1 |

Supple T2: Normalized Accent Counts

| **Language** | **count** |
| --- | --- |
| zulu | 141 |
| yoruba | 115 |
| swahili | 106 |
| igbo | 61 |
| hausa | 44 |
| xhosa | 37 |
| setswana | 32 |
| sesotho | 22 |
| pedi | 20 |
| twi | 17 |
| urhobo | 16 |
| tsonga | 14 |
| afrikaans | 13 |
| shona | 11 |
| luo | 6 |
| tagalog | 5 |
| ibibio | 8 |
| english | 5 |
| ebira | 5 |
| pidgin | 5 |
| ndebele | 5 |
| ewe | 4 |
| bekwarra | 4 |
| fante | 4 |
| venda | 4 |
| kikuyu | 4 |
| ijaw | 4 |
| bini | 4 |
| ibo | 3 |
| nembe | 3 |
| fulani | 3 |
| isoko | 3 |
| filipino | 3 |
| igala | 3 |
| jaba | 3 |
| kamba | 3 |
| kalabari | 3 |
| kwahu | 2 |
| efik | 2 |
| ika | 2 |
| tiv | 2 |
| ogoni | 2 |
| swati | 2 |
| bajju | 2 |
| ibani | 1 |
| patois | 1 |
| ga | 1 |
| urdu | 1 |
| dangme | 1 |
| izon | 1 |
| hindi | 1 |
| kalenjin | 1 |
| utugwang | 1 |
| evie | 1 |
| idoma | 1 |
| ukwani | 1 |
| ibra | 1 |
| edo | 1 |
| meru | 1 |
| tehl | 1 |
| bade | 1 |
| ham | 1 |
| atyap | 1 |
| adara | 1 |
| kanuri | 1 |
| french | 1 |
| eritrean | 1 |
| telugu | 1 |
| navajo | 1 |
| amo | 1 |
| limbum | 1 |
| luhya | 1 |
